# Effect of EBV Serology-based Screening Program on NPC Mortality: A Cluster Randomized Controlled Trial

**DOI:** 10.1101/2022.08.08.22278523

**Authors:** Wen-Jie Chen, Xia Yu, Yu-Qiang Lu, Ruth M. Pfeiffer, Wei Ling, Shang-Hang Xie, Zhi-Cong Wu, Xue-Qi Li, Yu-Ying Fan, Biao-Hua Wu, Kuang-Rong Wei, Hui-Lan Rao, Qi-Hong Huang, Xiang Guo, Ying Sun, Jun Ma, Qing Liu, Allan Hildesheim, Ming-Huang Hong, Yi-Xin Zeng, Ming-Fang Ji, Zhi-wei Liu, Su-Mei Cao

## Abstract

**Purpose:** To estimate the effectiveness of an Epstein–Barr virus (EBV) serology-based screening program to reduce nasopharyngeal carcinoma (NPC) mortality in a cluster randomized controlled trial in an NPC high-risk population.

**Methods:** Sixteen towns in Sihui and Zhongshan Cities, China were selected; eight were randomly allocated to the screening group and eight to the control group. Cantonese residents aged 30-69 years with no history of NPC were included January 1, 2008 to December 31, 2015. Residents in the screening towns were invited to undergo serum EBV VCA/EBNA1 IgA antibody tests; others received no intervention. Screening lasted through December 31, 2015; the population was followed through December 31, 2017.

**Results:** A total of 175,037 residents in the screening group and 184,526 residents in the control group were included. NPC incidence was similar in screening and control groups. A 28% reduction in NPC mortality was observed in the screening versus control arms in the overall study population (8.0 NPC deaths per 1000 person years versus 11.1; adjusted rate ratio [aRR]=0.72; 95% confidence interval=0.48-1.09; p=0.12). A stronger benefit was observed among individuals aged >50 (aRR=0.63; p=0.048) compared to those <50 (aRR 0.93; p=0.75). The reduction was increased among individuals from towns randomized to screening who participated in the screening program (aRR=0.38, p=0.001).

**Conclusion:** In this near 10-year trial, serology EBV VCA and EBNA1 IgA antibody testing led to a reduction in NPC mortality, particularly among individuals aged ⩾50. EBV antibody-based screening for NPC is effective at reducing NPC mortality in high-risk populations.

**Trial registration number:** NCT00941538

## Introduction

Nasopharyngeal carcinoma (NPC) has a unique geographic distribution, with approximately 85% of cases occurring in Asia^1^. NPC detection in its early stage is difficult because symptoms are non-specific; therefore, most cases are diagnosed at advanced stages, when 5-year overall survival is around 50% ^2^. In contrast, NPC 5-year overall survival approaches 95% when detected at early stages ^3^. Epstein-Barr virus (EBV) is a necessary cause for undifferentiated carcinoma (WHO type III), which accounts for approximately 95% of NPCs in endemic regions ^4,5^. This knowledge has led to the consideration of EBV-based strategies to reduce the NPC morbidity/mortality^6-11^.

Screening for NPC has been demonstrated to improve the early detection of NPC and cost-effectiveness models suggest such programs are highly cost-effective in endemic regions ^6,11-14^. However, the evidence supporting population-based screening remains incomplete. In particular, it is unknown whether NPC screening using EBV-based markers can reduce NPC-specific mortality in a screened population, largely due to lack of evidence from randomized controlled trials ^15^.

To address this important gap, we conducted a cluster randomized controlled trial to investigate the impact of EBV viral capsid antigen/nuclear antigen 1 (VCA/EBNA1) IgA antibody screening on NPC-specific mortality in two cities in southern China ^6^. The trial was initiated in 2008-2009 and included 16 towns for randomization in two cities (Sihui City and Zhongshan City) in Guangdong Province, China. Initial, interim analyses from the trial have shown that NPC cases detected through screening are typically early-stage ^6^ and that 4-year survival is improved for NPC patients in the screening group ^13^. Here, we present results from this randomized trial that evaluated the impact of EBV-VCA/EBNA IgA antibody screening on NPC-specific mortality over a 10-year period.

## Methods

### Study Design and Participants

The design of the screening trial has been described previously ^6,16^ and is provided in the **Study Protocol**. Briefly, sixteen towns in two cities (Sihui and Zhongshan) were selected as the cluster randomized units. We generated computerized random numbers and selected 8 towns as the screening group (Didou, Jianggu, Longpu, Luoyuan, Weizheng, Xiamao, Jingkou and Xiaolan) and the others (Chengzhong, Dasha, Dongcheng, Huangtian, Shigou, Zhenshan, Nantou and Dongfeng) as the control group. The trial focused on residents aged 30 to 59 in the initial phase (**study protocol Version 1.0 in Study Protocol**) and the eligible age range was broadened in 2012 to additionally include residents aged 60 to 69 years at time of randomization (**study protocol Version 2.0 in Study Protocol**) ^16^.

This screening trial (NCT00941538 Clinical Trials.gov) was approved by the Ethics Review Committee of the Sun Yat-sen University Cancer Center (SYSUCC, YP2009051).

### Recruitment

Baseline characteristics of the targeted population (totaling 359,563 residents 30 to 69 years of age) of Sihui and Zhongshan Cities at the time of screening initiation (2008 in Sihui and 2009 in Zhongshan) are shown in **Table 1**. All eligible residents in the screening towns were invited to participate by their village doctors through door-to-door visits, community education, and/or television advertisements. Written informed consent was obtained at the time of recruitment.

**Table 1.** Baseline and Follow-up Characteristics of Targeted Population.

| Variable | Control group | Screening group |  |  |
| --- | --- | --- | --- | --- |
|  |  | Total | Participants | Non-Participants |
| **Total No.** | 184,526 | 175,037 | 52,508 | 12,2529 |
| **Age at randomization** |  |  |  |  |
| Median age, years (IQR) | 44 (37-53) | 45 (38-55) | 44 (38-52) | 45 (38-57) |
| 30-39 | 66,907 (36.3%) | 54,575 (31.2%) | 16,335 (31.1%) | 38,240 (31.2%) |
| 40-49 | 55,752 (30.2%) | 53,877 (30.8%) | 19,153 (36.5%) | 34,724 (28.3%) |
| 50-59 | 41,698 (22.6%) | 44,096 (25.2%) | 15,473 (29.5%) | 28,623 (23.4%) |
| 60-69 | 20,169 (10.9%) | 22,489 (12.8%) | 1,547 (2.9%) | 20,942 (17.1%) |
| **Sex** |  |  |  |  |
| Male | 90,586 (49.1%) | 87,586 (50%) | 23,663 (45.1%) | 63,923 (52.2%) |
| Female | 93,940 (50.9%) | 87,451 (50%) | 28,845 (54.9%) | 58,606 (47.8%) |
| **City of residence** |  |  |  |  |
| Sihui City | 125,338 (67.9%) | 91,847 (52.5%) | 18,950 (36.1%) | 72,897 (59.5%) |
| Zhongshan City | 59,188 (32.1%) | 83,190 (47.5%) | 33,558 (63.9%) | 49,632 (40.5%) |
| **Town of residence** |  |  |  |  |
| Number of towns | 8 | 8 | n/a | n/a |
| Median number of residents<br>(range) | 20,331 (13,373-33,278) | 14,125 (9,017-20,339) | n/a | n/a |
| **In screening period (2008-2017)** |  |  |  |  |
| No. of deaths from any cause | 8,905 | 8,723 | 958 | 7765 |
| Cumulative all-cause mortality<br>(per 1000) (95% CI) <sup>a</sup> | 423.1 | 439.4 | 182.1 | 490.1 |
| Adjusted rate ratio of death from<br>any cause (95% CI) <sup>b</sup> | reference | 1.02 (0.91-1.16) | 0.46 (0.40-0.50) | 1.17 (1.05-1.30) |
| No. of NPCs | 656 | 632 | 189 | 443 |
| Cumulative NPC incidence (per<br>1000) (95% CI) <sup>a</sup> | 32.3 | 31.5 | 28.8 | 31.8 |
| Adjusted rate ratio of NPC<br>incidence (95% CI) <sup>b</sup> | reference | 0.97 (0.80-1.18) | 1.01 (0.80-1.28) | 0.96 (0.79-1.17) |

a Standardized to the China standard population in 2010
b Rate ratio calculated by Poisson regression, adjusting for cluster design (town), sex, age group, city of residence, and calendar year
Abbreviations: CI, confidence interval; IQR, Interquartile range; NPC, nasopharyngeal carcinoma

### Screening Methods and Management

Participants in the screening towns were offered serological tests for serum VCA IgA (EUROIMMUN AG, Lübeck, Germany) and EBNA1 IgA (Zhongshan Bio-Tech Company, Zhongshan, China) by ELISA ^17^. These markers were selected based on previous work that established an *a-priori* risk prediction algorithm ^17^, as follows: Logit *P* = -3.934 + 2.203×VCA/IgA + 4.797×EBNA/IgA. An NPC prediction score was calculated using this algorithm and used to established three NPC risk groups (high-risk, NPC score≥0.98; medium-risk, 0.65≤NPC score <0.98; low-risk, NPC score <0.65). Quality control for EBV antibody testing has been described previously. Briefly, we used pooled serum samples (calibrators) and the standards provided by the manufacturer to calibrate antibody levels across batches. The across-batch coefficients of variation (CVs) were 25% for VCA/IgA and 24% for EBNA1/IgA during the screening period.^16^

Participants categorized as “medium-risk” or “high-risk” were considered “seropositive” and invited to retesting annually; those categorized as low-risk were invited to rescreening at 5-yr intervals. Those with high-risk scores (NPC score ≥ 0.98) were referred to diagnostic work-up using fiberoptic endoscopy examination and nasopharyngeal biopsies if suspicious lesions were observed. During follow-up, those who seroreverted (from “medium-risk” or “high-risk” to “low-risk”) and had no noticeable lesion detected after three consecutive retests were shifted back into a 5-yr revisit schedule. Screening was conducted through December 31, 2015.

### Primary and Secondary End Points

The primary outcome of the trial was NPC-specific mortality. Secondary outcomes were the proportion of NPC cases diagnosed at stages I and II (i.e., early detection rate) and NPC survival. Clinical stage at diagnosis was defined according to the 2008 Staging System of China ^18^.

### Follow-up

Annual surveillance for NPC incidence/mortality, vital status, and immigration status for all the eligible subjects in the screening and control groups was conducted through linkage to Cancer, Death and Population Registries. The percentage of pathological confirmation of NPC cases was over 90% in the cancer registries from both cities included in our trial ^19,20^.

To ensure data accuracy, we conducted active follow-up by visiting each village/town to confirm the recorded information from local physicians through home visit and/or medial chart review. For each potential NPC case, available medical documentation was retrieved by an expert committee to review the tumor stage at diagnosis, treatment, and survival status. Medical charts were successfully retrieved for 1245 of the 1288 (96.7%) NPC cases identified during the study. Staging information was successfully obtained for 1149 (89.1% of 1288) of these NPC patients.

Among NPC cases, we observed 399 deaths (177 in the screening and 222 in the control groups; including 60 cases identified and confirmed through active follow-up only) during follow-up, of which 353 cases (88.5%; 90.5% in the control and 86.7% in the screening groups) were determined to have died due to NPC by the review committee blinded to screening group according to criteria shown in **Appendix 8 in Study Protocol**. Briefly, the criteria for reviewing the cause of death were based on presence of clinical evidence including metastases, local recurrence(s), and intervention-related death. The underlying cause of death was not NPC for the remaining NPC cases, of which 16 were determined to have died due to other cancers, 6 due to cardiovascular diseases, and 24 due to other causes. The last date of follow-up was December 31, 2017.

### Statistical Analysis

The primary analysis of the trial was a comparison of NPC-specific mortality between the screening and the control groups (main analysis: entire targeted population; sub-analyses: stratified by sex and age at randomization [<50 years and ≥50 years]), according to the intention-to-screen principle. We compared all-cause mortality and NPC incidence between the two groups to evaluate balance of characteristics across arms. Person time for each participant was the time from randomization to the date of first recorded NPC (used only for NPC incidence), death from NPC, death from any cause, or December 31, 2017, whichever came first. Event rates were defined as the ratio of the number of events over the person-years at risk for the event. Sex and age-group specific incidence rates were standardized to the 2010 standard Chinese population and then summed over all sex and age groups, towns and years of the trial. The test statistic *T* was the difference of the overall standardized rate in the screened towns and the unscreened towns. To accommodate the cluster randomized design in the p-value computation, we used a permutation test where we randomly assigned 8 towns to the screened group and 8 to the unscreened group and repeated the computation of *T*. The p-values for *T* were based on this permutation distribution. In addition to the permutation test, for each outcome, we also fitted Poisson regression models to the observed number of events including the log of the person years as an offset, and adjusted for age-group, sex and screening assignment. Town was included using a random effect.

To further evaluate the impact of serum EBV antibody screening, we divided residents in the screening towns into those who participated in the EBV antibody tests (hereinafter referred to as “participants”) and those who did not (hereinafter referred to as “non-participants”).

NPC-specific cumulative incidence and mortality curves were estimated using the Nelson-Aalen estimate. Overall and stage-specific survival curves among patients with NPC were estimated using the Kaplan-Meier method. Compared with NPC patients in the control group, p*-*values for differences between survival curves were obtained by log-rank tests.

Statistical analyses were performed using SAS version 9.4 (SAS Institute Inc, Cary, North Carolina) and R statistical software 4.1.2. All statistical tests were 2-sided, and p*-*values <0.05 were considered statistically significant.

## Results

### Baseline and Follow-up Characteristics of the Targeted Population

The screening group (8 towns) included 175,037 residents at the time of randomization and the control group (8 towns) included 184,526 residents (excluding 2060 residents who voluntarily attended the screening tests) (**Figure 1**). In 2007 (before the trial initiation), the NPC incidence rate in the screening group was slightly lower than in the control group (18.0 per 100,000 person-years vs. 21.9) but the NPC mortality rate was slightly higher (14.3 per 100,000 person-years vs. 12.2). Baseline characteristics did not differ significantly between the two groups, except for the city of residence (by design) (**Table 1**). At randomization, the median age was 44 years in the control group (interquartile range [IQR], 37 to 53) and 45 years in the screening group (IQR, 38 to 55).

**Figure 1.**
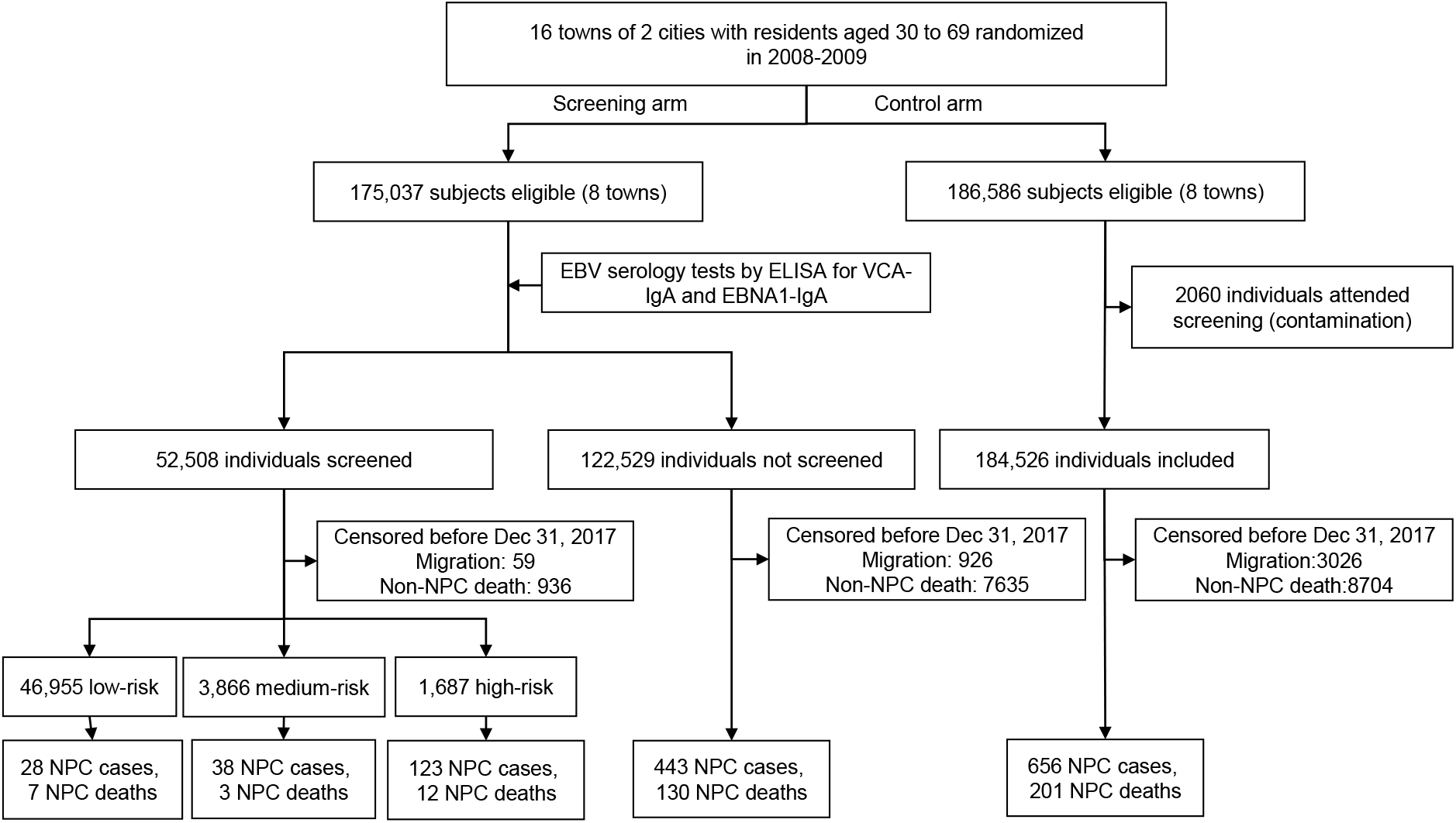
Trial flow diagram. Abbreviations: EBV, Epstein-Barr virus; EBNA1, EBV nuclear antigen 1; NPC, nasopharyngeal carcinoma; VCA, EBV viral capsid antigen

We observed 8,723 deaths with 1,624,304 person years of follow-up in the screening group and 8,905 with 1,731,895 person years in the control group (**Table 1; Supplementary Table 1** lists the specific causes of death in the screening and control groups). Since NPC constitutes ∼2% of total deaths in the population, we did not expect our screening trial would have had an impact on the all-cause mortality. As expected, cumulative all-cause mortality at the end of follow-up was similar in the two groups. The standardized all-cause mortality (standardized to the China population in 2010) was 439.4 per 1000 person-years in the screening group and 423.1 per 1000 person-years in the control group (adjusted rate ratio [aRR] comparing screening versus control, 1.02; 95% CI=0.91-1.16, p=0.70). **Supplementary Figure 1A** shows the cumulative all-cause mortality for the screening and control groups.

Similarly, we did not expect screening would have detected non-aggressive NPC; therefore, the NPC incidences between the screening and control groups would be comparable. As expected, the standardized cumulative NPC incidence was 31.5 per 1000 person-years in the screening group and 32.3 per 1000 person-years in the control group (aRR comparing screening versus control, 0.97; 95% CI=0.80-1.18, p=0.77). **Supplementary Figure 1B** shows the cumulative incidence of NPC by group.

In the screening group, 52,508 (30.0% of 175,037) residents participated in the serum EBV antibody test. Details on the screening visits by baseline serological risk group are shown in **Supplementary Table 2**. The overall compliance rate of nasopharyngeal endoscopic examination and/or biopsies among the high-risk individuals classified according to baseline results was 65.9% (1110/1687). Compared to the control group, participants were more likely to be older, female, and live in Zhongshan City (**Table 1**). Fewer deaths were observed in participants than in control group during the screening period (2008-2017; **Supplementary Figure 2A)**, with the aRR of death from any cause of 0.46 (95%CI: 0.40 to 0.50, p=0.0042). In contrast, participants and control subjects were similar with respect to NPC incidence (aRR=1.01; 95%CI=0.80 to 1.28; p=0.91). The cumulative numbers of NPC cases over time in the screening group (stratified by participation) and control group are shown in **Supplementary Figure 2B**.

### NPC-specific Mortality

At the end of follow-up, 152 individuals in the screening group and 201 in the control group had died from NPC. The standardized cumulative NPC mortality rate was 8.0 per 1000 person-years in the screening group and 11.1 in the control group, with the absolute rate difference of 3.0 per 1000 person-years (95%CI, -2.0 to 8.0 per 1000 person-years). The aRR for death from NPC (screening group vs. control group) was 0.72 (95% CI, 0.48 to 1.09, p=0.12, **Table 2**). The cumulative numbers of NPC deaths over time are shown in **Figure 2A**.

**Table 2.** Cumulative Nasopharyngeal Carcinoma (NPC)-specific Mortality (2008-2017)

| **Group** | **No. of deaths**<br>**from NPC** | **Person-**<br>**years** | **Cumulative NPC-**<br>**mortality rate**<br>**(per 1000) <sup>a</sup>** | **Adjusted rate ratio**<br>**(95%CI) <sup>b</sup>** | **p-value <sup>c</sup>** |
| --- | --- | --- | --- | --- | --- |
| Control group | 201 | 1,731,895 | 11.1 | reference |  |
| Screening group | 152 | 1,624,304 | 8.0 | 0.72 (0.48-1.09) | 0.120 |
| Participants | 22 | 488,506 | 5.8 | 0.38 (0.22-0.68) | 0.001 |
| Non-participants | 130 | 1,135,798 | 10.6 | 0.83 (0.55-1.26) | 0.380 |

<sup>a</sup> Standardized to the China standard population in 2010
<sup>b</sup> Rate ratio calculated by Poisson regression, adjusting for cluster design (town), sex, age group, city of residence, and calendar year
<sup>c</sup> *P*-values calculated based on Poisson regression

**Figure 2.**
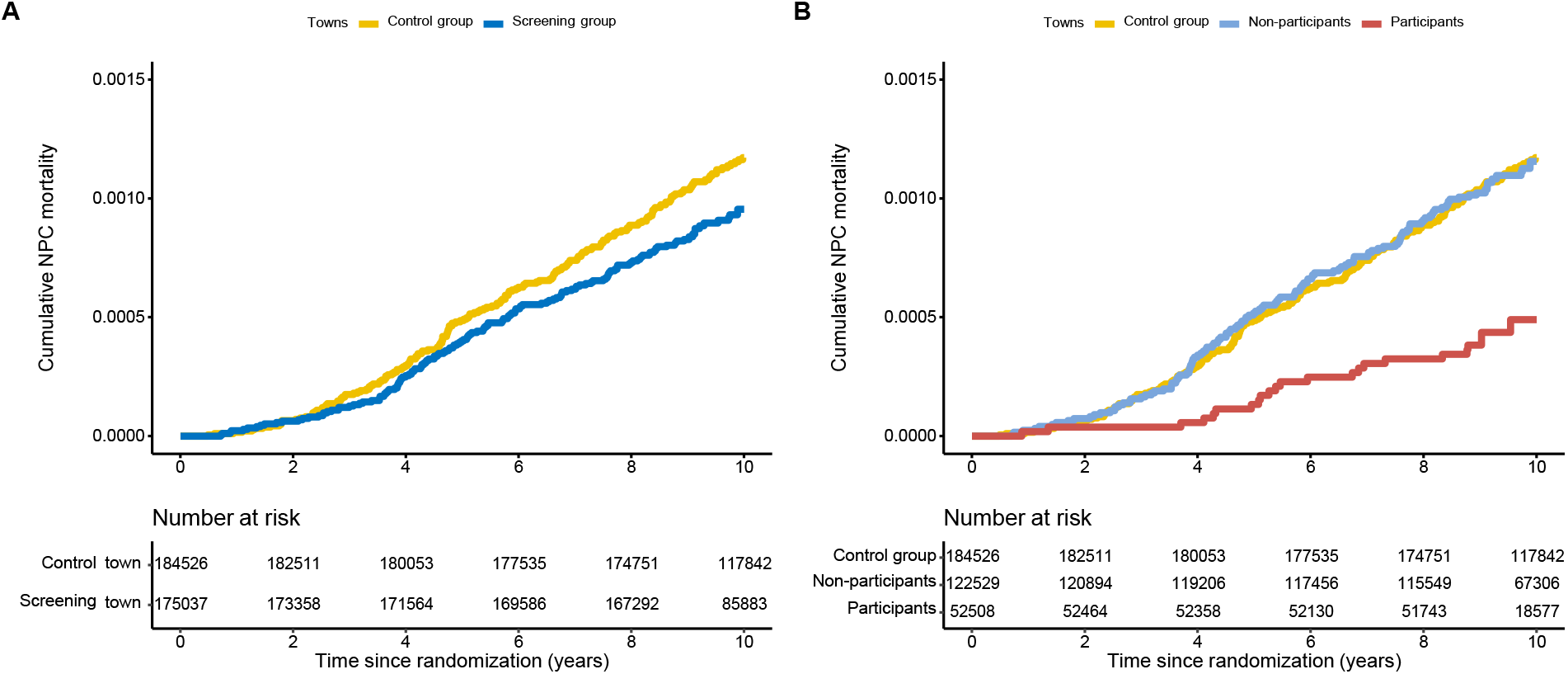
Cumulative numbers of nasopharyngeal carcinoma (NPC) deaths during follow-up, stratified by A) screening towns vs. control towns; B) participants and non-participants in the screening towns vs. control towns

In the screening group, serum EBV antibody tests were significantly associated with reduced NPC-specific mortality among participants. The standardized cumulative NPC mortality rate was

5.8 per 1000 person-years among participants, with the absolute rate difference of 5.3 per 1000 person-years (95%CI, -2.0 to 13.0 per 1000 person-years) compared with the control group. The aRR estimate for death from NPC among participants was 0.38 (95% CI, 0.22 to 0.68; p= 0.001), compared with the control group (**Table 2** and **Figure 2B**).

### Subgroup Analyses for NPC-specific Mortality

**Figure 3**. shows the person-years of residents who underwent randomization, the numbers of those who died from NPC, and the rate ratio for the subgroups of sex and age group (<50 and ≥50 years), according to the screening group (overall and among participants) and control group. Results for all-cause mortality and NPC incidence are shown in **Supplementary Figures 3 and 4**.

**Figure 3.**
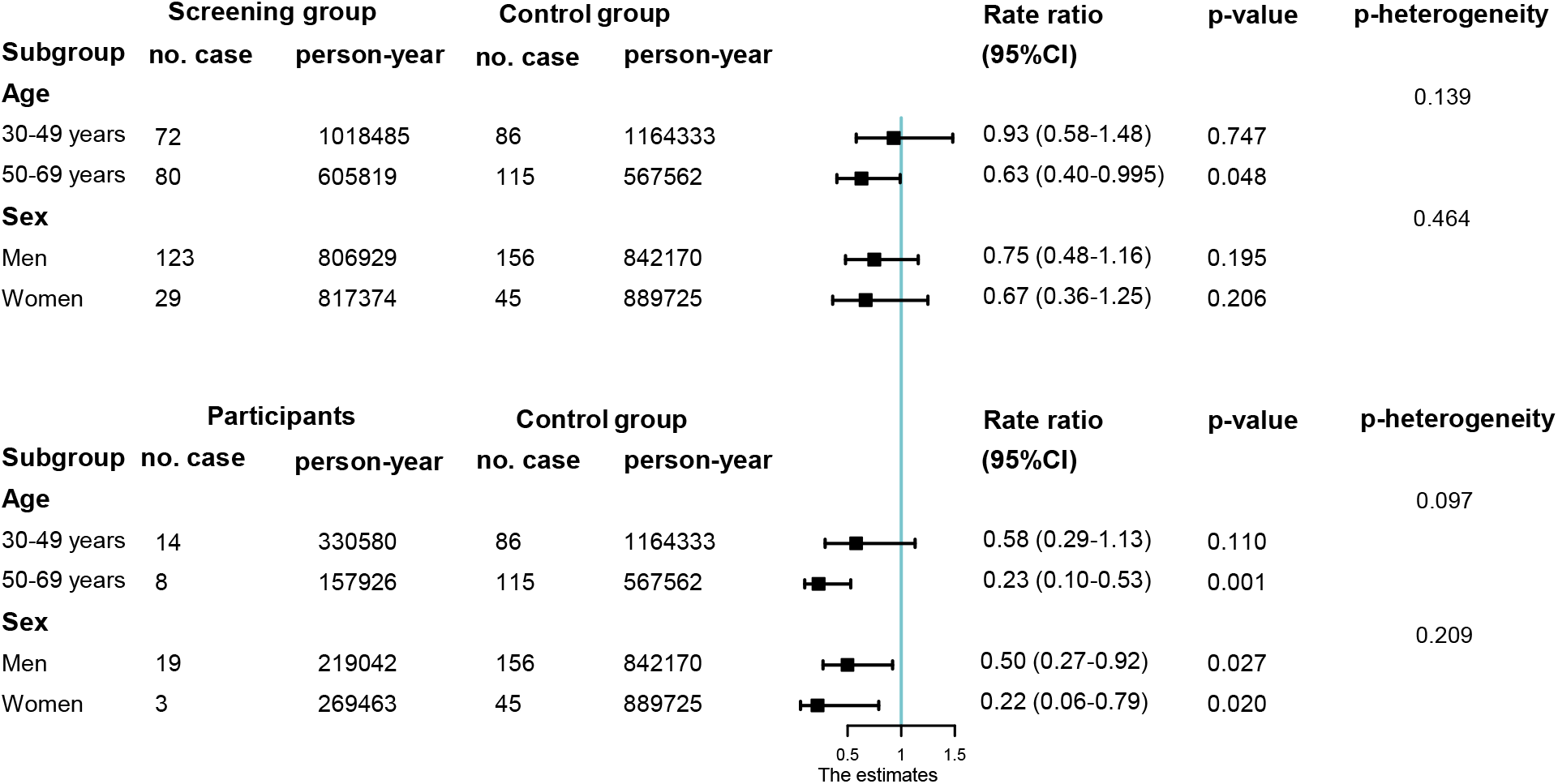
Adjusted rate ratio of death from nasopharyngeal carcinoma (NPC) by sex and age group (<50 and ≥50 years). A) screening group vs. control group; B) participants in the screening group vs. control group. Rate ratios were calculated by Poisson regression, adjusting for cluster design (town), sex, age group, city of residence, and calendar year

Compared with the control group, the effect of screening on NPC-specific mortality was similar for both sexes. However, screening of residents aged 50+ years had a stronger effect on NPC-specific mortality. For example, the aRR of death from NPC was 0.93 (95% CI, 0.58 to 1.48; p=0.75) for the younger age group vs. 0.63 (95% CI, 0.40 to 0.995, p=0.048) for the older age group (p-heterogeneity=0.14) (**Figure 3A**). The heterogeneity of reduction in NPC-specific mortality due to age was larger when participants were compared to the control group, with the aRR of death from NPC of 0.58 (95% CI, 0.29 to 1.13) for the younger age group and 0.23 (95% CI, 0.10 to 0.53) for the older age group (p-heterogeneity=0.10) (**Figure 3B**).

### NPC Characteristics

**Table 3** presents characteristics of NPC cases by screening group, and for screening participants and non-participants. The histological subtypes were similar across groups; most cancers were undifferentiated nonkeratinizing carcinoma (WHO type III; 96.0 % in the control group, and 93.7% in the screening group). The early detection rate in the screening group was significantly higher than in the control group (31.0% in Stages I and II vs. 22.6%, p=0.001). In the screening group, the early detection rate in participants was highest, with 55.0% being in early stages (i.e., stages I and II). There was no significant difference in clinical stages in cases detected from non-participants in the screening and control groups (20.0% vs. 22.6%, p=0.29). Cases detected in the screening group (and among screening participants) had a lower probability of death compared with those from the control group, and cases detected from participants also had better survival than non-participants and control groups (**Supplementary Table 3 and Supplementary Figures 5 and 6**).

**Table 3.**
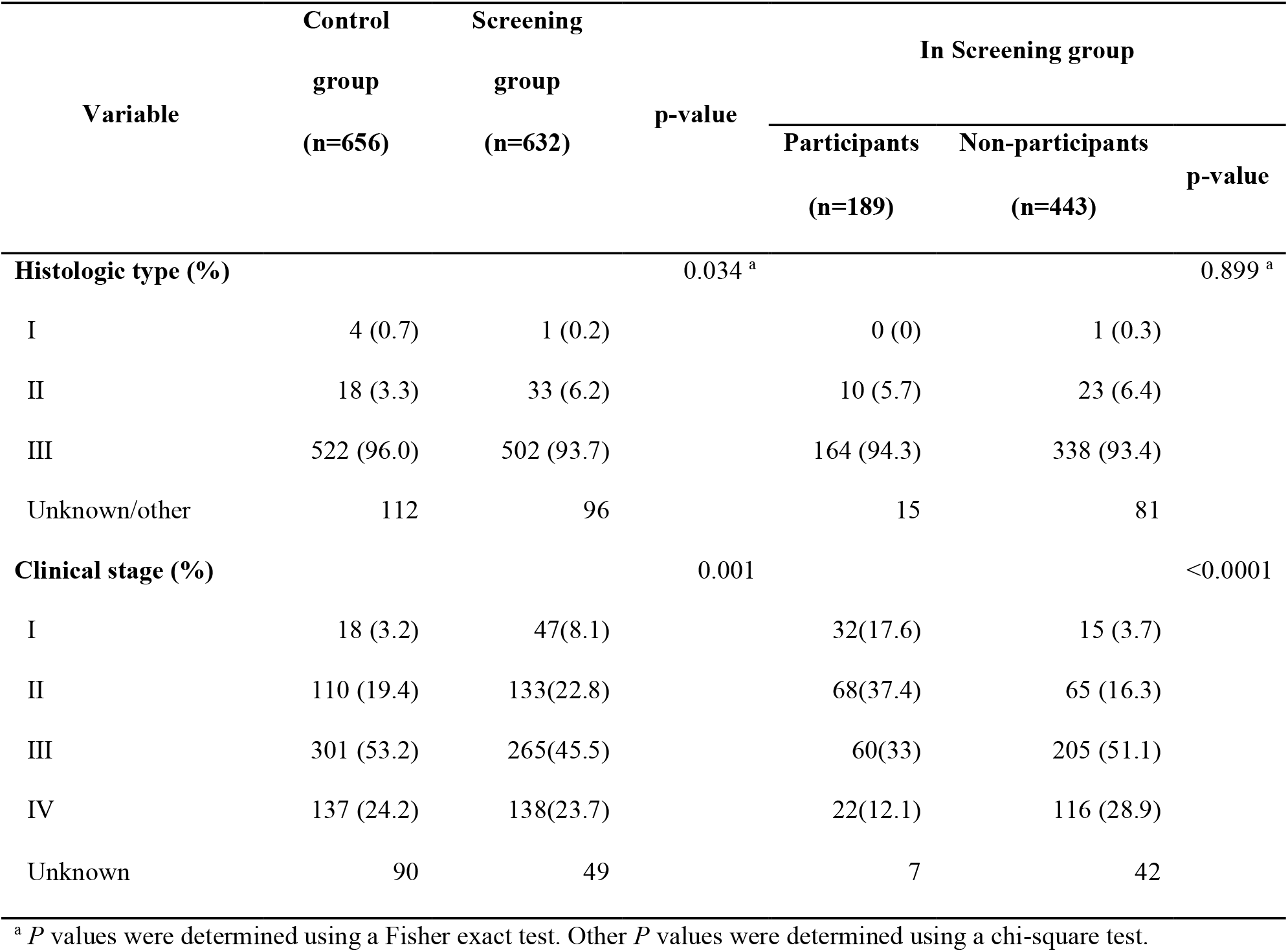
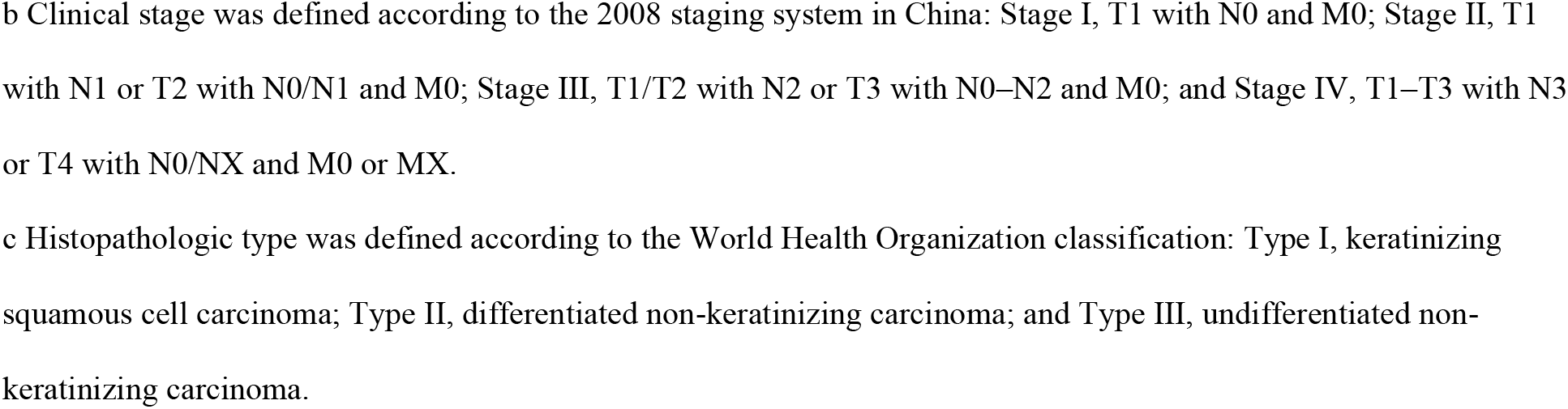
Nasopharyngeal Carcinoma (NPC) Histologic Type and Stage in the Targeted Population.

## Discussion

Growing evidence supports the use of EBV-based markers for NPC screening. However, whether NPC screening can be effectively translated into public health programs that reduce mortality associated with NPC remains unclear. In a recent consensus statement on EBV-based screening for NPC^21^, evaluation of the impact of screening on NPC mortality was identified as an important future research objective. Understanding whether NPC screening leads to the identification of less aggressive forms of NPC was also important for future work. In the present report, we address both issues for the first time. Results from our cluster-randomized clinical trial with nearly 10 years of follow-up show that, compared to individuals from towns randomized to routine care (control group), those randomized to EBV VCA/EBNA IgA antibody screening experienced a near 30% reduction in NPC mortality, an effect that was more pronounced (near 40%) and statistically significant among individuals aged ≥50 years. Furthermore, within towns randomized to screening, individuals who agreed to be screened had an over 60% reduction in mortality compared to individuals from control towns.

Consistent with the observed reduction in mortality, we also observed considerable “down-staging” of disease stage at diagnosis among NPC cases diagnosed among individuals who underwent screening compared to those who did not, with 55.0% of NPC cases diagnosed at stages I/II. These results are in accordance with our previous report from an interim analysis conducted in one of our participating centers (i.e., Zhongshan City) at 4.5 years of follow-up ^13^ and extend them by reporting results from all participating centers, followed for a longer time period (near 10 years), and including a broader age group (30-69 years).

The ideal age to start screening for NPC has not been established and the current screening recommendations are primarily based on the age-specific incidence rate in high-risk populations, which starts to increase steeply at age 30 in both men and women and exhibits a single peak at approximately ages 55-59 years in the area where our screening trial was conducted ^22,23^. As noted above, we observed a stronger reduction in mortality among individuals screened at 50-69 years, suggesting the impact of screening can be maximized by focusing screening efforts on this age group. Whether the higher NPC mortality reduction observed among those 50-69 years in our trial is due to the higher NPC incidence in this age group compared to younger individuals or to the fact that older individuals are more likely to comply with screening remains to be determined.

Screening programs aimed at early detection and mortality reduction need to guard against the possibility of inadvertently concomitantly leading to overdiagnosis of non-lethal, non-aggressive forms of the disease being screened for ^24-26^. By demonstrating that overall incidence of NPC was similar across trial arms, our study suggests that NPC-specific mortality can be reduced through screening without a concomitant increase in the overdiagnosis of previously unrecognized forms of NPC that would not have come to the attention of the medical system in the absence of screening. This observation supports that the mortality benefit of NPC screening was not at the expense of a substantial harm of excess detection of additional non-aggressive cancers that would never have been diagnosed. Our finding further suggests and is consistent with the clinical observation that this tumor is prone to invasion and metastasis in the natural state, and that the overdiagnosis of indolent tumors commonly observed for other epithelial cancers is not a common phenomenon in NPC ^27^.

Only 30% of eligible participants in our trial chose to avail themselves of screening, and among those who did and who screened positive, only 66% complied with the recommended clinical follow-up and biopsy. Our results show that the reduction in NPC-specific morality was considerably larger among those who participated in the screening program, suggesting that EBV antibody screening for NPC has the potential of achieving larger NPC-specific mortality reduction than those reported herein should successful efforts be made to increase participation and compliance with screening programs. However, it is noted that participants who complied with screening in our trial were healthier than non-participants as indicated by the lower all causes mortality among participants during study period. Therefore, it is currently uncertain how much the additional NPC mortality reduction observed among screening participants compared to non-participants in the towns randomized to screening is attributable to their compliance with screening versus their overall increased health status. Future studies should aim to obtain additional information regarding the characteristics of non-participants and the reasons for non-compliance with screening recommendations to better understand this phenomenon and to help develop strategies to increase participation in NPC screening programs.

Strengths of this study include the large sample size and cluster randomized design that included a contemporaneous control group. The fact that mortality reduction was observed in both cities included in the study suggests the feasibility of reproducibly implementing the same screening protocol across centers and the generalizability of our results in NPC high-incidence settings. However, one important limitation as discussed above, is the low participation rate among individuals randomized to the screening group (i.e., 30%). In the future, more flexible screening implementation should be considered to maximize participation, particularly among the highly mobile population in southern China. Second, the cluster randomization approach used for our trial is more prone to bias than trials that randomize at the individual level ^28,29^. Third, the question of whether a reduction in morbidity among NPC cases can be achieved through screening was not addressed in the present trial. While our observation that NPC cases detected through screening tended to be diagnosed at an earlier stage (and therefore require less aggressive treatment) is suggestive that long-term morbidity can be minimized through screening, future studies should aim to formally address this question.

In nearly 10 years of follow-up, our NPC screening trial in a high-risk population demonstrated that EBV VCA/EBNA IgA antibody testing can lead to a reduction in NPC mortality, particularly among those who are screened between the ages of 50-69. Furthermore, our results suggest that efforts will be needed to maximize compliance with screening recommendations to maximize the impact of screening programs. Additional research is warranted to understand the benefit of screening at different ages and to better quantify morbidity reduction associated with screening programs.

## Data Availability

All data produced in the present work are contained in the manuscript.

## Acknowledgment

This work was supported by National Key R&D Program of China (2020YFC1316905); National Natural Science Foundation of China (81872700, 82073625); Eleventh National Science and Technology Support Program of China, National Key Research and Development Program of China (2017YFC0907100, 2016YFC0902000); Sun Yat-Sen University Clinical Research 5010 Program (2013012); Intramural Research Program of National Cancer Institute (NCI), USA.

## Figure Legend

**Supplementary Figure 1.**
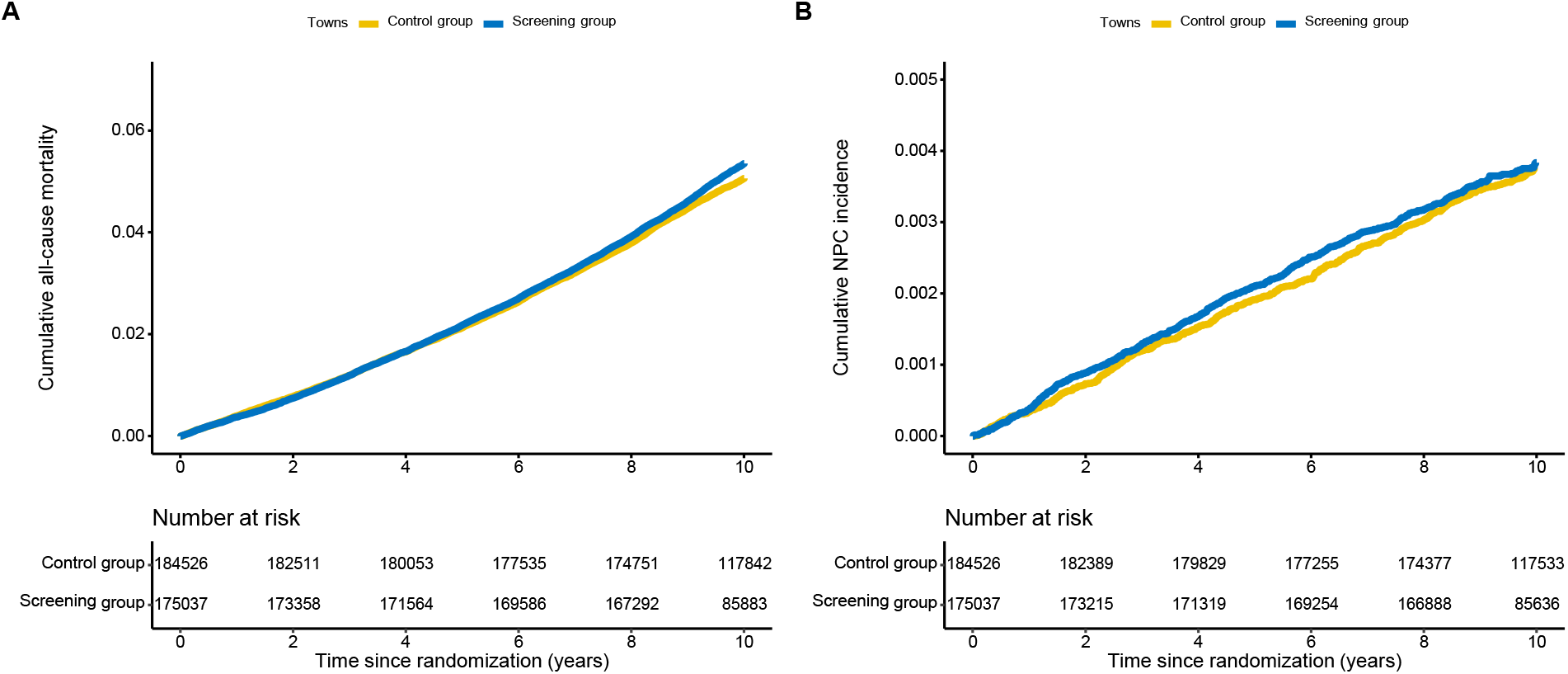
Cumulative all-cause mortality and incidence of nasopharyngeal carcinoma (NPC) by screening group and control group. A) Cumulative all-cause mortality; B) Cumulative incidence of NPC

**Supplementary Figure 2.**
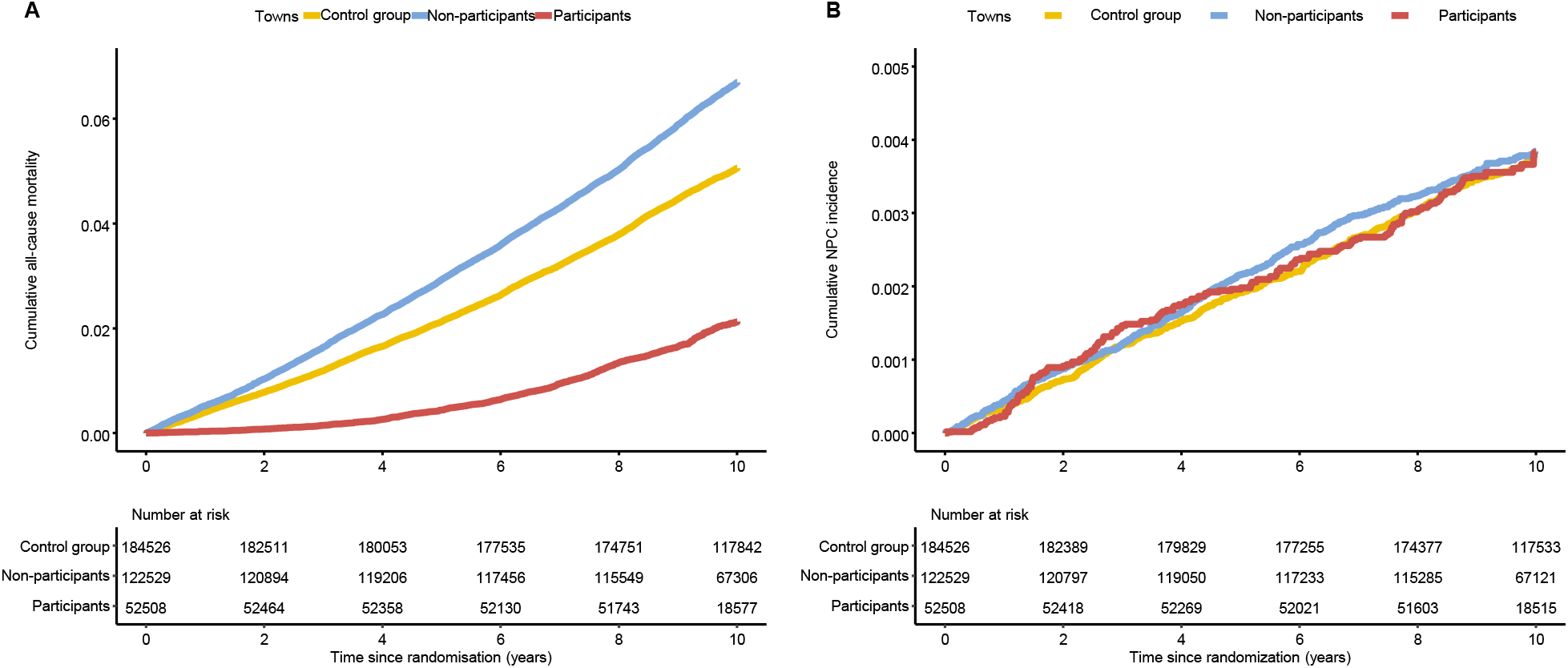
Cumulative all-cause mortality and incidence of nasopharyngeal carcinoma (NPC) by participation status in the screening group, compared to control group. A) all-cause mortality; incidence of NPC.

**Supplementary Figure 3.**
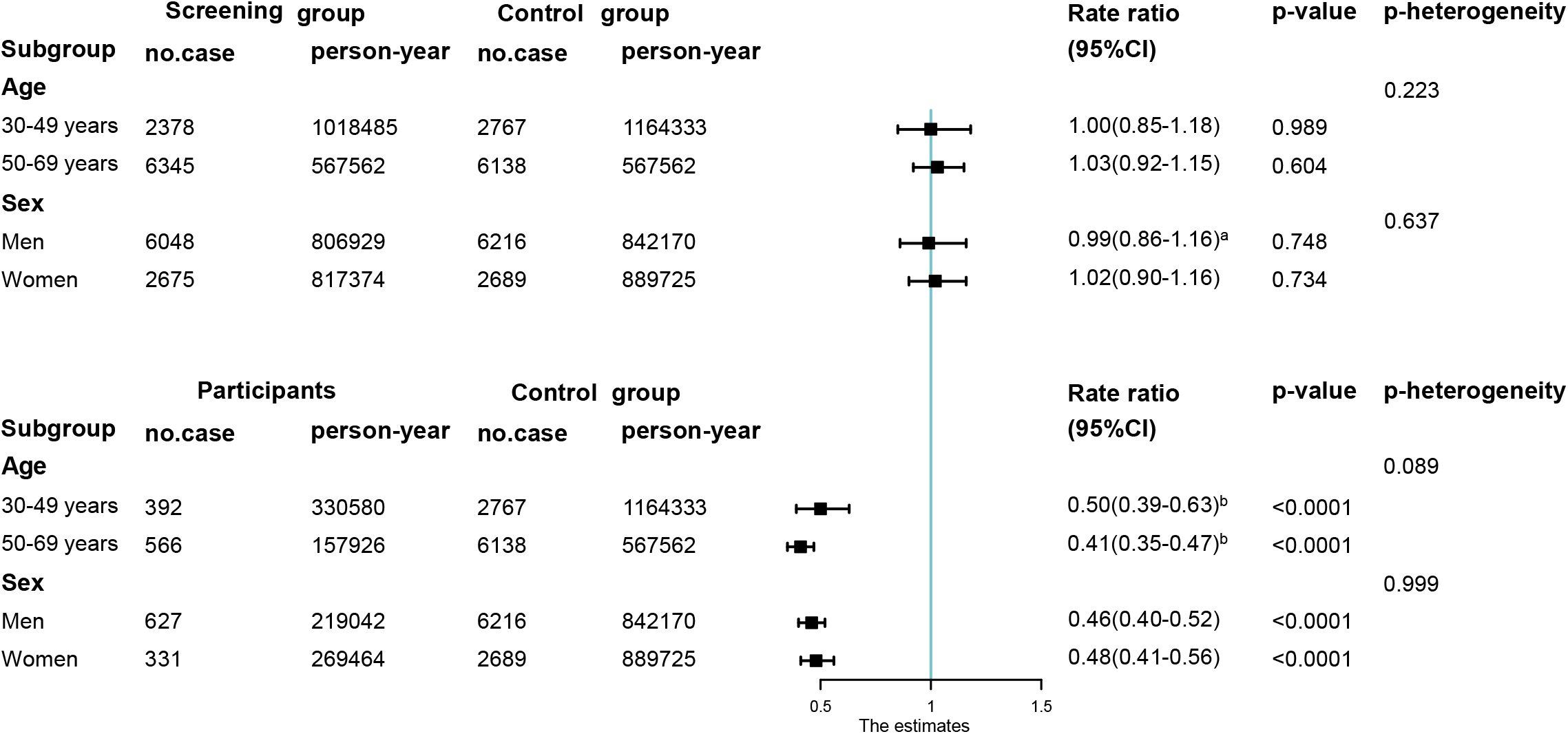
Forest plots showing the adjusted rate ratios for all-cause mortality by sex and age. A) screening group vs. control group; B) participants in the screening group vs. control group. *a. Models were adjusted for calendar year as a continuous variable b. Models were not adjusted for age and year

**Supplementary Figure 4.**
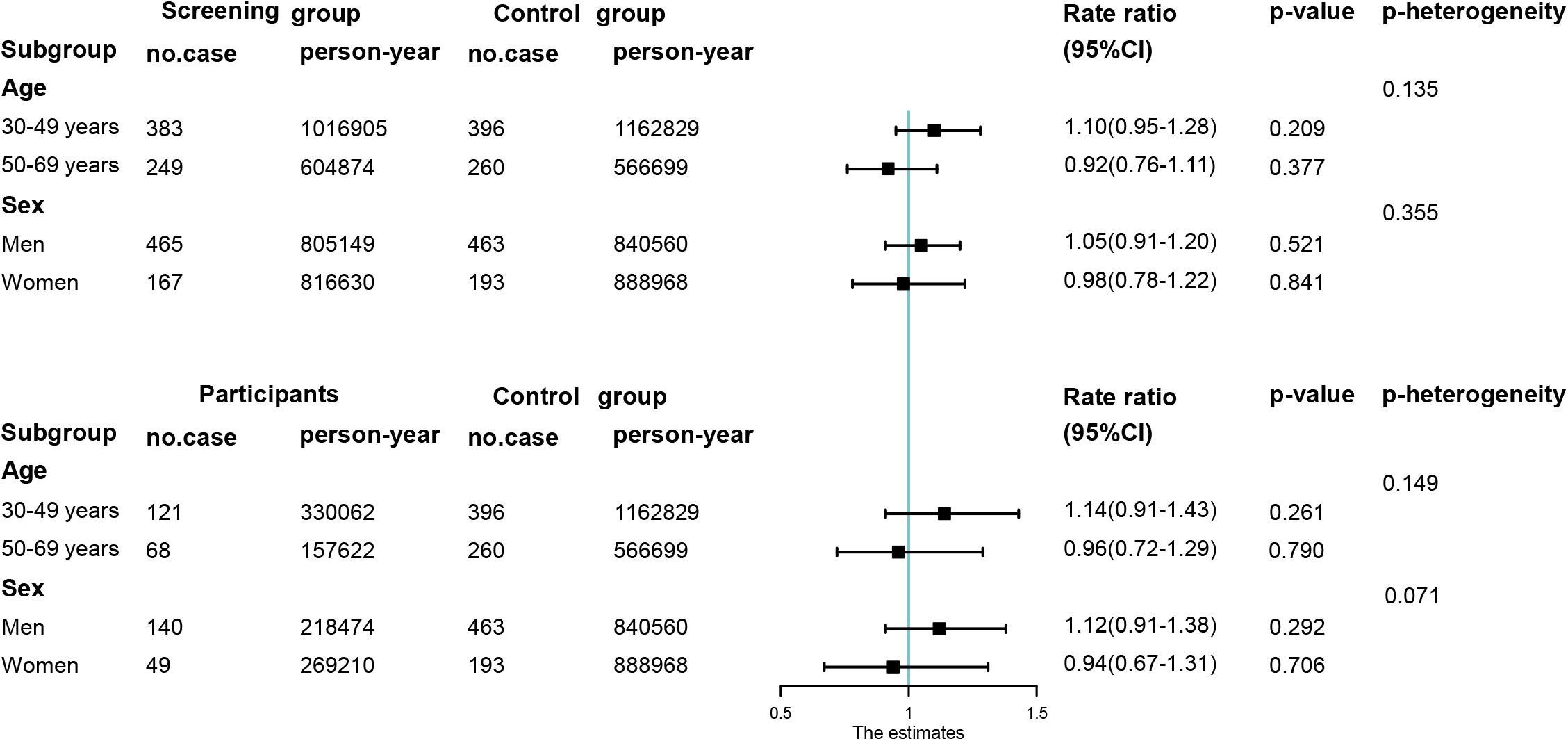
Forest plots showing the adjusted rate ratios for incidence of nasopharyngeal carcinoma (NPC) by sex and age. A) screening group vs. control group; B) participants in the screening group vs. control group. All models were fitted without a random effect of towns due to zero counts in some strata.

**Supplementary Figure 5.**
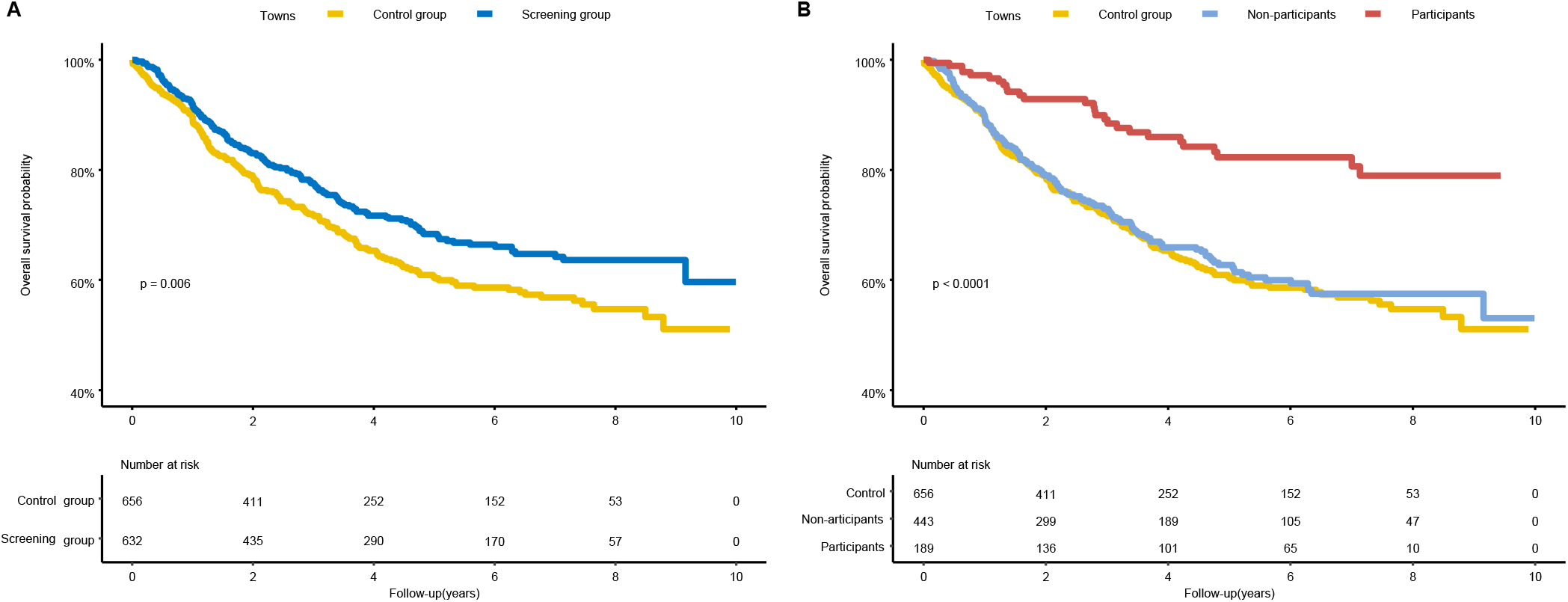
Survival curves of patients diagnosed with nasopharyngeal carcinoma (NPC) among A) screening group vs. control group; B) participants and non-participants in the screening group vs. control group. *P*-values were obtained by log-rank tests.

**Supplementary Figure 6.**
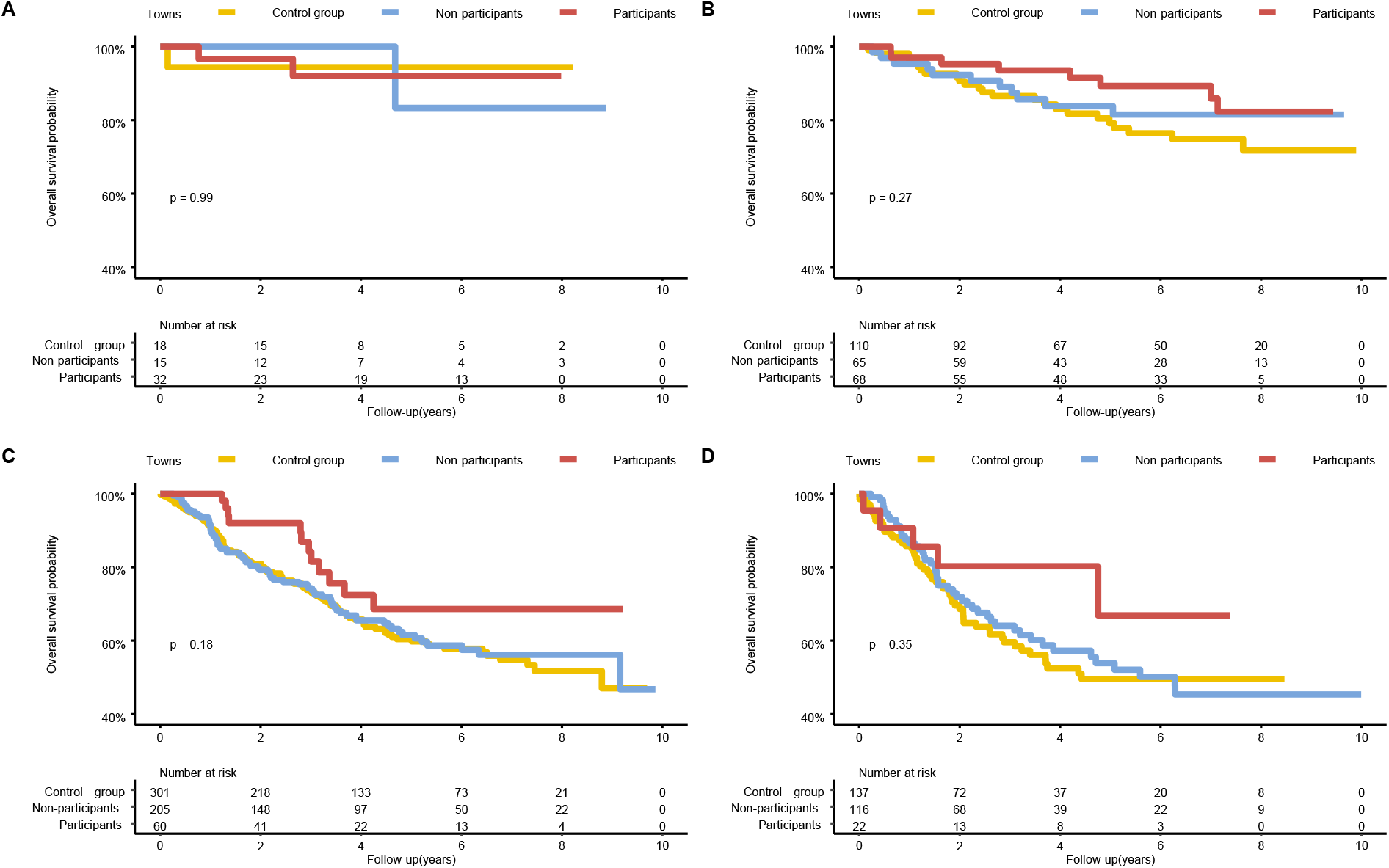
Survival curves of patients diagnosed with nasopharyngeal carcinoma (NPC) by clinical stage. A) Stage I; B) stage II; C) Stage III; D) stage IV. *P*-values were obtained by log-rank tests. Clinical stage was defined according to the 2008 staging system in China: Stage I, T1 with N0 and M0; Stage II, T1 with N1 or T2 with N0/N1 and M0; Stage III, T1/T2 with N2 or T3 with N0–N2 and M0; and Stage IV, T1–T3 with N3 or T4 with N0/NX and M0 or MX.

**Supplementary Table 1.** Cause of Death of Deceased Population Excluding Nasopharyngeal Carcinoma Death.

| Cause of Death | Control group | Screening group |  |  |
| --- | --- | --- | --- | --- |
|  |  | Total | Participants | Non-participants |
| **Total No.** | 8704 | 8571 | 936 | 7635 |
| Infectious disease | 74(0.9%) | 92(1.1%) | 4(0.4%) | 88(1.2%) |
| Cancer (excluding NPC) | 2840(32.6%) | 2569(30%) | 315(33.7%) | 2254(29.5%) |
| Endocrine and metabolic diseases or immune disorders | 164(1.9%) | 159(1.9%) | 11(1.2%) | 148(1.9%) |
| Nervous system diseases | 79(0.9%) | 93(1.1%) | 12(1.3%) | 81(1.1%) |
| Circulatory disease | 2476(28.4%) | 2283(26.6%) | 186(19.9%) | 2097(27.5%) |
| Respiratory disease | 685(7.9%) | 595(6.9%) | 73(7.8%) | 522(6.8%) |
| Digestive disease | 427(4.9%) | 452(5.3%) | 81(8.7%) | 371(4.9%) |
| Accidental | 490(5.6%) | 465(5.4%) | 41(4.4%) | 424(5.6%) |
| Urogenital disease | 181(2.1%) | 233(2.7%) | 28(3%) | 205(2.7%) |
| Other | 1288(14.8%) | 1630(19%) | 185(19.8%) | 1445(18.9%) |

**Supplementary Table 2.**
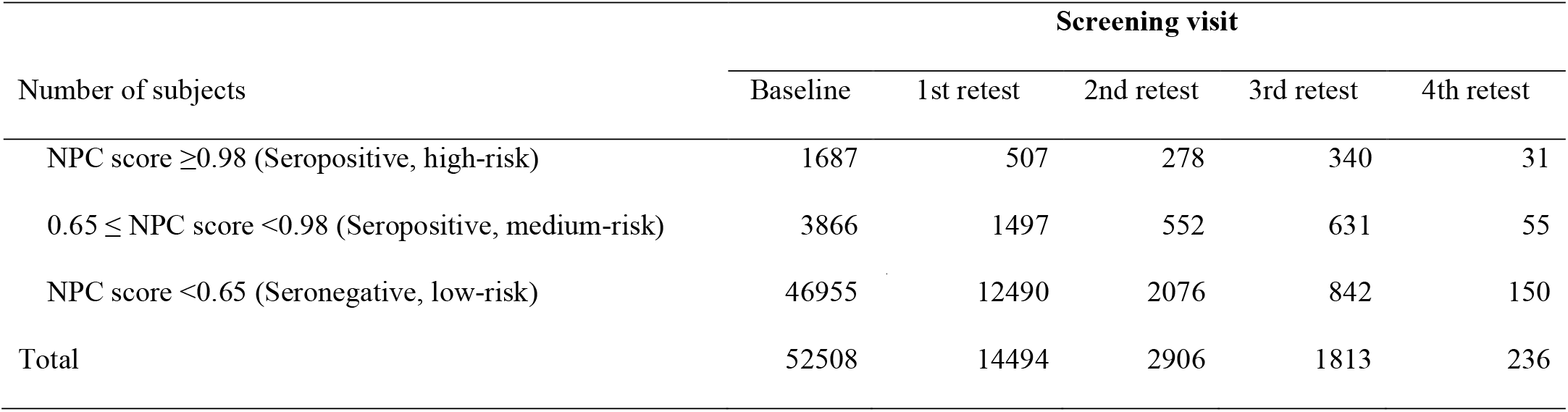
Number of Participants by Baseline Serological Group in Relation to Screening Visit.

| Number of subjects | Screening visit |  |  |  |  |
| --- | --- | --- | --- | --- | --- |
|  | Baseline | 1st retest | 2nd retest | 3rd retest | 4th retest |
| NPC score $\geq 0.98$ (Seropositive, high-risk) | 1687 | 507 | 278 | 340 | 31 |
| $0.65 \leq$ NPC score $< 0.98$ (Seropositive, medium-risk) | 3866 | 1497 | 552 | 631 | 55 |
| NPC score $< 0.65$ (Seronegative, low-risk) | 46955 | 12490 | 2076 | 842 | 150 |
| Total | 52508 | 14494 | 2906 | 1813 | 236 |

**Supplementary Table 3.** Overall Survival of NPC Patients by Clinical Stage.

| **Group** | **5-year survival rate**<br>**(95% CI)** | **8-year survival rate**<br>**(95% CI)** |
| --- | --- | --- |
| **Overall** |  |  |
| Control group | 0.61 (0.56-0.65) | 0.55 (0.50-0.60) |
| Screening group | 0.68 (0.64-0.73) | 0.64 (0.59-0.68) |
| Non-participants | 0.63 (0.58-0.68) | 0.58 (0.52-0.63) |
| Participants | 0.82 (0.76-0.89) | 0.79 (0.72-0.87) |
| **Early Stage (Stages I and II)** |  |  |
| Control group | 0.81 (0.74-0.89) | 0.74 (0.65-0.85) |
| Screening group | 0.88 (0.82-0.93) | 0.83 (0.77-0.91) |
| Non-participants | 0.84 (0.76-0.93) | 0.82 (0.74-0.92) |
| Participants | 0.90 (0.84-0.97) | 0.85 (0.76-0.95) |
| **Late Stage (Stages III and IV)** |  |  |
| Control group | 0.57 (0.52-0.63) | 0.51 (0.45-0.58) |
| Screening group | 0.61 (0.56-0.67) | 0.55 (0.49-0.62) |
| Non-participants | 0.59 (0.53-0.65) | 0.52 (0.46-0.60) |
| Participants | 0.68 (0.56-0.83) | 0.68 (0.56-0.83) |

